# Facilitators and barriers to the implementation of the biobehavioral survey among incarcerated individuals and correctional personnel in Mozambique, 2021

**DOI:** 10.1101/2023.07.04.23292209

**Authors:** Carlos Botão, Ana Mutola, Samuel Nuvunga, Auria Banze, Rachid Muleia, Makini Boothe, Cynthia Semá Baltazar

## Abstract

Mozambique implemented in 2021 a formative assessment in 22 prisons to identify the operational and logistical needs for the second round of the Biobehavioral Survey (BBS). We discussed barriers and facilitators that could be anticipated in conducting BBS in prisons, in preparation for implementation, structured interviews with key informants with directors and other staff were administered. The data were cleaned and analyzed in Microsoft Excel, and the categorical variables were summarized by means of simple frequencies and percentages. In most prisons the actual prison capacity far exceeds the theoretical capacity, 40.9% have a theoretical capacity of ≥ 50 inmates, 81.8% have inmates who exceed their theoretical capacity. In the country half of the prisons receive only male inmates, and only one female, 54.5% of the prisons visited have inmates under 18 years of age, 72.7% of the prisons had a private space available for the survey, the penitentiary establishments have the physical space for study; ensuring the safety of staff within the facilities; involvement of correctional officers and a clinical focal point. However, barriers such as time management due to prison opening hours, prison laws, restrictions, or permits for research may change without notice due to security, lockdowns, riots, or other situations that may hinder the implementation of research. Currently, little is known about the implementation of a BBS in a correctional environment and only a few barriers can be anticipated, for Mozambique’s context, these challenges and obstacles can be overcome through clear communication and collaboration with officials at all levels.

## Background

It is estimated that there are, at any given time, nearly eleven million people held in prison settings around the world, and about 668,000 of those are in the sub-Saharan African region (UNAIDS, 2021). At the global level, the prevalence of Human Immunodeficiency Virus (HIV), tuberculosis (TB), hepatitis (B and C), and other Sexually Transmitted Infections (STI) are reportedly as ben higher among inmates when compared with the general population (Dianatinasab et al., 2018). Several structural factors inherent to the prison conditions, including overcrowding, human rights violations, lack of access to prevention and treatment services, as well as behavioral factors, such as sexual activity between men (including forced sex), drug use (including injecting drugs), and tattooing or the practices of shaving are all associated with HIV transmission among prisoners.

Although data are very limited, the Sub-Saharan African region has one of the highest HIV prevalence rates among inmates (Azbel et al., 2016a). Penitentiary establishments and other closed settings are one of the most neglected in the regions’ responses to addressing HIV in low- and middle-income countries. The rigid and poor prisons infrastructure sometimes makes it difficult to implement preventive programs against the transmission and spread of the disease. Furthermore, when this population transitions back to the community, they can continue to spread disease to the general population since they did not receive the required treatment while incarcerated (Azbel et al., 2016b).

The sudden outbreak of the COVID-19 pandemic highlighted human rights concerns for people deprived of their liberty such as prisoners. The overcrowded and confined conditions, poorly ventilated infrastructure, unsanitary conditions, as well the justice system and legal representation amplified their susceptibility to high rates of COVID-19 infections and mortality in incarcerated settings (P et al., 2020; Wang et al., 2020).

Due to the “*revolving-door”* effect, wherein inmates, prison staff, and prison visitors move in and out of the prison, there is great potential for the transmission of diseases to home communities and to other incarceration facilities, thus further contributing to the spread of these communicable pathogens across the population (Meyer et al., 2014, Telisinghe et al., 2016).

Frequent prisoners transfer and the cycle of recidivism can also impede the continuity of care for those undergoing treatment for communicable diseases, particularly for those requiring long-term treatment such as HIV and TB. These disrupted treatment cycles can contribute to increased pathogen resistance at the population level (Biadglegne et al, 2015).

In Mozambique, it is often reported that the prison health service does not have access to the equipment and medicines needed to provide adequate medical care. The quality of health care that prisoners have access to was generally assessed as inadequate. With regard to inmates with physical disabilities, there is little or no provision made for your needs. The death rate of prisoner’s in certain prisons was considered excessive. This aspects would require further investigation into whether these incidents are related to poor access to health, care and / or conditions of detention that could be addressed (REFORMAR, 2018).

The HIV epidemic in the country is a critical public health issue with an estimated HIV of 13.2% among adults over the age of 15, one of the highest in the world (Ministério da Saúde (MISAU) et al., 2015; Semá Baltazar et al., 2021). In order to control the HIV epidemic, Mozambique is working to increase utilization and access to prevention, testing and treatment programs for several key populations [for example, female sex workers (FSW), men who have sex with men (MSM) and people who inject drugs (PWID), who are generally at higher risk of HIV infection than the general population. These populations also face a number of legal and social barriers that affect their use and access to health services, as well as increase their vulnerability to HIV. Incarcerated people are recognized by the Mozambican government as a key population in the National Strategic Plan on HIV/AIDS (Council of ministers, 2015).

According to Mozambique’s National Strategic HIV and AIDS Response Plan (2015-2019), the objectives for key populations are to ensure that this have access to quality, evidence-based health services in order to contribute to an effective response to the HIV epidemic in Mozambique (PEN IV, 2015). In response, the National Program for STI and HIV/AIDS Control developed the National Guidelines for the Integration of HIV/AIDS Prevention, Care and Treatment Services for Key Populations in the Health Sector (PEN IV, 2015). These guidelines present a set of ten (10) strategic interventions to address the specific prevention and treatment needs of key populations. At present, the implementation of the comprehensive service pack is focused on FSW and MSM, while efforst has not yet been released for PWID and prisoners due to logistical issues.

The results curried out in 2013, as established in the assumptions that justified the study, it was expected that the results of the HIV, STIs and TB were more than twice the prevalence in the general population, or in prisoners as well as in penitentiary agents. The report indicated that 24% (22.3-25.9) of inmates in Mozambican penitentiary establishments were infected with HIV; 16% (14.5-17.5) were infected with treponema pallidum and 1.5% (0.9-2.0) for the Tuberculosis bacillus. The same indicates that 18.5% (13.6-24.8) of the penitentiary agents of Mozambican penitentiary establishments were infected with HIV and 9.7% (6.0-14) are infected by trepomena pallidum. None penitentiary agents was positive for the bacillus of Tuberculosis in the year in which the study took place. The report concludes that HIV prevalence among prisoners in penitentiary establishments is high, but lower than expected. The presence of STI symptoms is associated with HIV infection in encarcerated people (Ministry of Justice, 2013). The prevalence of HIV among penitentiary agents in penitentiary establishments is high and very above the prevalence in the general population with men being the most affected (UNODC, 2013).

The Correctional System in Mozambique faces yet numerous difficulties related to infrastructure, allocation of providers for health, social, and other services, and the existence of infectious diseases such as HIV and other STIs, TB, malaria, skin diseases, etc. Among the different diseases, communicable disease is the primary problem in the prison environment, and transmission can continue when individuals return to their communities. In addition to needing to care for the health and well-being of the incarcerated population, they require special attention because their release and re-integrate back into the community can have repercussions on the dynamics of transmission and his/her reinfection for HIV, Syphilis and others STIs (Augusto et al., 2017; Vaz et al., 1996).

As a way of following up on the surveillance activities for this type of population, the INS in collaboration with its partners carried out in 2021 the first round of the BSS in the incarcerated population and penitentiary agents with a view to monitoring the progress achieved by interventions aimed at the prevention of HIV infection, provision of care and treatment services in this population.

This kind of survey in general can advise the Ministries of Health and of Justice and the Constitutional and Religious Affairs to join efforts as a way to create and harmonize policies that promote and improve health in the correctional system. Acting in a more integrated perspective can contribute to the construction of a correctional health system closer relied on human rights and on more equitable access to health in the Correctional Facilities of Mozambique.

Before the implementation of the survey itself, a formative evaluation was carried out in all provinces of the country to identify the operational and logistical needs for carrying out the main phase of the survey in each location, to deepen knowledge about the characteristics of the incarcerated population and correctional officers, as well as to assess the conditions of infrastructure and other specific information related to this study.

In this paper, we provide information related to the barriers and facilitators in the implementation of an ethically and scientifically sound BBS survey among the prisoner population in Mozambique, to assist future prevention and treatment efforts.

## Methodology

A formative assessment was conducted in 22 penitentiary establishments across all 11 provinces (2 establishments per province) between May and August 2021. The penitentiary establishments were selected using a methodology of systematic probability proportional to size with implicit stratification. these establishments size was specified as the square root of the penitentiary establishments population and province was used as the stratification variable. This approach was used to maximize the geographic representation of the selected establishments. The size of the incarcerated population (at least 50) and the organizational structure (health center, security conditions, infrastructure conditions to accommodate the survey team, visual and auditive privace) of each establishment were taken into account for inclusion in the sample. All penitentiaries stated that they have an updated list containing the number of inmates that might be available if requested in advance for the purpose of random and anonymous recruitment (ensuring confidentiality about the description of the identity of the participants). All penitentiary establishments were chosen in consultation with Correctional Service from the Ministry of Justice.

### Study population

The formative assessment was conducted among 22 penitentiary establishments directors, staff and some incarcerated people; they were all over 18 years old. They were identified as key informants since they possessed knowledge of the working mechanisms of each establishment, the characteristics, behaviors, attitudes, and practices of inmates and correctional officers, as well as the type of services existing or not of the penitentiary establishment. All individuals provided written informed consent for the participation in semi-structured interviews.

### Data collection

A semi-structured interview guide was developed with questions about the functioning of penitentiary establishments, types of existing assistance services (health, social action, social reintegration, etc.), capacity of inmates, opening hours, among others. The interviews were conducted by trained interviewers and data collected using Tablets and open-source ODK platform.

In addition to the data collection carried out with key informants, in-person observations were carried out at each sampled establishment. The observations were done to primarily assess the establishment intake process, confirm the presence of health centers, evaluate the conditions of the health centers, and potential locations amenable to the implementation of the BBS survey and to ensure confidentiality. If so, make sure this is mentioned in the methods section as well as storage data and materials. Before the implementation of the formative assessment, the data collection team members received training, including an overview of the incarcerated population as well as correctional officers, ethical aspects in research involving human beings, vulnerabilities, and standardized operational procedures for data collection. All information captured from the questionnaires and observations were sent to the central server housed at the Data Management Unit of INS in Marracuene Village. The investigators closely supervised data collection and conducted some of the interviews.

### Data analysis

Descriptive data were cleaned and analyzed using Microsoft Excel, and categorical variables were summarized using simple frequencies and percentages. The qualitative data were categorized and analyzed thematically.

## Assessment Results

A total of 22 penitentiary establishment were assessed (two in each province). In most establishment, the actual prison capacity far exceeds the theoretical capacity. Many establishments (40.9%) have a theoretical capacity of 50-100 inmates, however based on data proved most of this (81.8%) have inmates that exceed their theoretical capacity.

Half (50%) of the establishments only house male inmates, and only one is specific to female inmates (located in the capital city, Maputo City). More than half (54.5%) of 22 establishments visited have inmates under 18 years of age. About three quarters (72.7%) of the establishments had a private space available to conduct the survey (table I).

**Table 1.** Description of sampled prisons, Mozambique 2021.

| Variables | n (N=22) | % |
| --- | --- | --- |
| **Current Population size** |  |  |
| 50 -100 inmates | 9 | 40.9 |
| 101-250 inmates | 8 | 36.4 |
| 251-500 inmates | 1 | 4.5 |
| >500 inmates | 4 | 18.2 |
| **Nr of Prisons that exceed their capacity limit** | 18 | 81.8 |
| **Type of prison** |  |  |
| Male-only | 11 | 50.0 |
| Female-only | 1 | 4.5 |
| Mixed | 10 | 45.5 |
| **Incarceration of inmates under the age of 18** | 12 | 54.5 |
| **Nr of prisons that participated in a previous survey/study round** | 6 | 27.3 |
| **Infrastructure available to conduct the survey** |  |  |
| Meeting rooms | 4 | 18,2 |
| Open field | 8 | 36.4 |
| Health unit/observation room | 7 | 31.8 |
| Permanence | 1 | 4.5 |
| No infrastructure available | 1 | 4.5 |
| **Had a private space available to conduct the survey** | 16 | 72.7 |

### Facilitating Factors or Study Implementation

#### Coordination and Partnership

Several researchers report challenges to get the permission to conduct research with incarcerated individuals (Fox et al., 2011). We found and identify as facilitators the fact that a close collaboration established with the Ministry of the Interior, which oversees the Department of Corrections, is essential to successful survey implementation. All required authority figures within the Ministry of the Interior provided their approval and co-signed an authorization letter describing the general methodology, taking into consideration establishment-specific operations and programmers such as operation hours, eating schedule, workouts and others. As the Department of Corrections is a hierarchical institution at Ministry of Interior and therefore all approvals must first be granted by the respective Directors.

Most establishment were willing to provide physical spaces – such as meeting rooms, medical care offices, or the atrium – where the team can perform data collection activities and store non-sensitive study materials. Privacy during the data collection process was prioritized and the majority (72.7%) of the prisons sampled contained a suitable setting to conduct the study.

#### Safety and Security of Research Team

The research team’s manifestation of interest in carrying out the research in the establishments led the directors of each establishment to guarantee the safety of the team within the facilities requiring guards to escort study teams, transport inmates to the study area and provide extra security. However, in all prisons sampled, the availability of mechanisms and security devices was guaranteed in these terms. Such availability would reduce the number of inmates outside their cells, the use of correctional officers escorting study teams, and the need for guards to protect team members during BBS implementation. The involvement of both key correctional officials and a clinical focal point is crucial for conducting any public health research in prisons. The survey team (including investigators and interviewers) should be accompanied in designated areas (where the survey will be conducted) in the establishments (frame 1).

**Frame 1.** Summary of facilitating factors to BBS implementation, Mozambique 2021

| **Theme** | **Summary** |
| --- | --- |
| **Official authorization** | *Ministry of Defense provided the authorization and all the required support to access the prisons facilities.* |
| **Private space to conduct the survey** | *The majority were able to identify spaces with privacy where surveys can be implemented: reception, private room to conduct questionnaire and sample collection.* |
| **Samples and material storage** | *Most have a place for the conservation of samples collected during study hours and store materials.* |
| **Infrastructure for study team** | *Most provide access to changing rooms for the team, pantry and create pathways for the team's entry into and exit from the prison during the study.* |
| **Acceptance of study devices and tools** | *All prisons will allow study staff to enter carrying laptops, cellphones, and tablets for purposes of data collection.* |
| **Safety and security** | *In all prisons, the availability of mechanisms and security devices was guaranteed.* |

### Possible barriers for study implementation

One of the major concerns with conducting a BBS in establishments is time management due to the establishments operating hours. It was agreed that the survey could only be conducted within the operating hours of state institutions (from 7:30 am to 3:30 pm). Gaining access into the establishment and proceeding through the necessary security checks can take up to thirty minutes for each study team member. The team also needs to consider the inmates’ mealtimes. Timing can impact the number of individuals that can be interviewed and the length of time that can be spent with them. Efficient planning will be key in successful BBS implementation.

Establishment laws, restrictions or permissions for research can change unannounced due to security, lockdowns, rebellions, or other situations that may disrupt survey implementation. Investigators need to be aware of this possibility and it is imperative to have members on the study team who are familiar with the system and functioning of prisons facilities.

The COVID-19 pandemic has undeniably changed the manner in which survey procedures are carried out and the impact extends to penitentiary establishments. Although incarcerated, prisoners are not isolated from the world and are therefore extremely vulnerable to SARS-CoV-2, since they congregate in areas with poor ventilation. In order to mitigate SARS-CoV-2 infection, a series of measures at national level were adopted. All establishment employ protocols to prevent the spread of COVID-19 in the penitentiary establishment. This fact plays an important role in the implementation of this survey, contributing equally to the safety of the members of the research team, as well as the participants. It should be noted due to current COVID-19 prevention protocols, the establishments limit the number of inmates who can be outside of their cells at a given time. The majority (54.5%) of penitentiary establishments allowed 5-10 inmates out of their cells at any time. About a quarter (27.3%) of penitentiary establishments only allowed 1-5 inmates out of their cells at a time. How inmates are recruited to be BBS participants, the limitation on the number of inmates allowed out of their cells directly impacts BBS implementation, specifically the amount of time full participation would be required.

The penitentiary establishment is a self-contained environment in which activities are strictly regulated and monitored. According to research norms and principles in Mozambique, including IRB, there is a need for additional protections for vulnerable populations in general and incarcerated persons specifically. The informed consent documentation states that participation of an incarcerated person in research is voluntary and should not affect parole or correctional programming decisions. Unlike other surveys where participants can call the principal investigator at any time to request clarifications or report adverse events, in this environment the participants do not have as much autonomy. Participants must therefore receive enough and understandable information to make a voluntarily informed decision. Another important aspect is participation *bias*. The arrival of the research team in the closed environment can, in itself, affect incarcerated perceptions of the institutions in which they live. Prisoners may be more inclined to participate in a study simply because it would involve getting the attention of an interviewer. On the other hand, incarcerated people may be suspicious of researchers. Establishing trust to collect accurate information is very important and crucial.

About three quarters (77.3%) of the penitentiary establishments sampled reported having an internal health facility to provide care to inmates and other people in the establishment. However, the majority of respondents (directors and/or their substitutes) referred to mobile clinics that travel to penitentiary establishments to provide healthcare services to inmates in need. On other occasions, inmates are taken to the reference health facility for more private care. It was also mentioned that some prisons receive weekly visits from a health professional.

About 23% of penitentiary establishments do not have a community-based organization that works with inmates on the establishment. The community-based organizations play a significant role on outreach and HIV/AINDS peer education.

## Conclusion

Incarcerated persons are regarded as a vulnerable population for research study purposes, including the BBS. The opportunity to conduct a BBS should ultimately result in benefits for the inmates, for example by improving conditions or access to health services for vulnerable populations.

Currently, little is known about the implementation of a BBS in a correctional setting and only some barriers can be anticipated. However, in the current context of Mozambique, these challenges and obstacles can be overcome through clear communication and collaboration with officials at every level, from the individual prison to the Ministry of the Interior.

## Data Availability

All data produced in the present study are available upon reasonable request to the authors
All data produced in the present work are contained in the manuscript

## Declaration

The protocol approved the Mozambique National Committee on Bioethics for Health. For all participants written informed consent was obtained.

## Availability of data and materials

The dataset analyzed for the current study are fully available at the Mozambique National Institute of Health (INS) data repository for researchers who meet the criteria for access to confidential data. Data are from the IBBS study’s whose authors may be contacted through: www.ins.gov.mz.

## Competing interests

The authors declare that they have no competing interests

## Funding

This paper used data from research supported by the Global Fund trough the Mozambique Ministry of Health

## Authors contributions

All authors have read and approved the final version of the manuscript.

## Acknowledgments

The authors would like to thank Jordan McOwen and Neha Kamat for his critical revision.

## Notes

### Competing Interest Statement

The authors have declared no competing interest.

### Author Declarations

The protocol approved by the Mozambique National Committee on Bioethics for Health. For all participants written informed consent was obtained.

